# Values and preferences of the general population in Indonesia in relation to COVID-19 self-testing: A cross-sectional survey

**DOI:** 10.1101/2022.01.23.22269718

**Authors:** Caroline Thomas, Sonjelle Shilton, Catherine Thomas, Deepshikha Batheja, Srishti Goel, Claudius Mone Iye, Elena Ivanova, Guillermo Z. Martínez-Pérez

**Author notes:** Corresponding author: Sonjelle Shilton.

## Abstract

**Objectives:** Innovative diagnostics are essential to assist members of the general population become active agents of case detection. In Indonesia, a country with an over-burdened healthcare system, individuals could use self-tests for SARS-CoV-2 to determine their COVID-19 status. To assess the acceptability of SARS-CoV-2 self-testing among the general population in Indonesia, a cross-sectional, population-based survey was conducted in mid-2021 in Jakarta and the provinces of Banten and North Sulawesi.

**Methods:** This was a survey that approached respondents in >600 randomly selected street-points in the three study geographies. A 35-item questionnaire was used to collect data on key variables, such as willingness to use and to pay for a SARS-CoV-2 self-test and likely actions following a positive result. Bivariate and multivariate regression analyses were performed.

**Results:** Of 630 respondents, (318 were female), 14% knew about COVID-19 self-testing, while 62.7% agreed with the concept of people being able to self-test at home, unassisted, for COVID-19. If self-tests were available in Indonesia, >60% of respondents would use them if they felt it necessary and would undertake regular self-testing e.g., weekly if recommended. Upon receiving a positive self-test result, most respondents would communicate it (86.03%), request post-test counseling (80.79%), self-isolate (97.46%), and/or warn their close contacts (*n*=570, 90.48%).

**Conclusions:** SARS-CoV-2 self-testing would be acceptable to a majority of the Indonesian public, to learn whether they have COVID-19. Self-testing could contribute to an over-burdened healthcare system by helping COVID-19-infected people become agents of change in epidemiological surveillance of SARS-CoV-2 in their communities.

## INTRODUCTION

In December 2019, the first case of a person infected by the novel coronavirus SARS-CoV-2 was reported in Wuhan, China [1]. The World Health Organization (WHO) declared this disease to be a pandemic in March 2020 [2] by which time Indonesia was among the most affected countries, with an 8.9% case fatality rate by the end of March 2020 [3]. As of 11 January 2022, this nation of more than 270 million inhabitants had dealt with over 4,266,649 cases, with close to 150,000 deaths attributable to COVID-19 [4].

In an effort to control the pandemic in Indonesia, country-wide, screening interventions to detect new cases of COVID-19 were implemented [5]. Testing for SARS-CoV-2 infection in laboratories and clinical settings was scaled-up to include individuals with COVID-19-compatible symptoms and close contacts of confirmed cases [5, 6]. However, systematic screening for COVID-19 cases requires a significant investment in human resources and diagnostic technology that Indonesia cannot afford without compromising the quality of healthcare for patients.

Currently, Indonesian pharmacies dispense saliva-based rapid antigen tests for COVID-19 to those wishing to check their status without requesting testing at a Jaminan Kesehatan Nasional (i.e., national health insurance) facility [7, 8]. The concept of self-testing is not novel; diagnostics for the home detection of infectious diseases such as human immunodeficiency virus (HIV) have been on the market since the mid-1990s in the USA [9]. Self-testing devices for private, home- or work-based detection of SARS-CoV-2 infection could be promoted to increase case detection while allowing the healthcare system to prioritize its diagnostic services for persons with COVID-19-compatible symptoms and their close contacts [10]. However, to ensure that SARS-CoV-2 self-testing in Indonesia has an impact on case detection, reduction of clinic workloads, and reduction of COVID-19 morbimortality, society-grounded strategies are necessary to introduce this innovation to the public in such a way that isolation, contact tracing, and effective requests for confirmatory testing and further clinical care can occur following any positive result. Therefore, information about the Indonesian general public’s values around SARS-CoV-2 self-testing is required. To address this, a survey was conducted to assess communities’ attitudes toward self-testing. Other objectives of this survey were to understand the drivers of self-testing acceptability, willingness to pay for self-testing, and likely actions taken upon a positive result. Here, we describe the survey findings that could be useful to inform regulatory and healthcare decision-making in relation to SARS-CoV-2 self-testing in Indonesia.

## METHODS

### Design, population, and sites

This was a cross-sectional, population-based survey conducted between July and August 2021. The survey population was the general population of three geographies in Indonesia: the capital city of Jakarta, and the provinces of Banten (Java) and North Sulawesi (Celebes). The eligibility criteria were that participants had to be aged ≥18 years, willing to provide informed consent, and without symptoms compatible with COVID-19 disease.

Jakarta was selected to represent urban dwellers’ views, while Banten and North Sulawesi were selected to represent rural dwellers’ views. Specifically, North Sulawesi was selected to represent the non-Jawa and non-Moslem minority populations of Indonesia. To understand whether differences based on geographical location influenced the acceptability of SARS-CoV-2 self-testing, sample size calculations were performed separately for each site. It was estimated that 196 or more respondents at each of the three sites, for a total of 588, would be necessary to have a confidence level of 95%, so that the real value (acceptability of COVID-19 self-testing) was within ±7% of the measured value.

### Sampling and recruitment

A multi-staged sampling process was applied. First, one site per geography was selected, with the boundary of each site defined using Google MyMaps®. Once defined, the site maps were divided into 40 areas of similar width, which were numbered from 1 to 40. Second, using a random number generator (RANDOM.ORG®), the three lists of 40 areas were randomly reordered and the first 14 areas in the newly arranged lists were selected as recruitment areas. Third, using the same random number generator, the three lists of 14 areas were reordered again to determine the sequence that the surveyors would follow when visiting each area during the survey. Fourth, in each of the 14 areas, 21 randomly selected street-points were manually marked and were where the study staff would be stationed to recruit participants.

When conducting the survey pairs of surveyors arrived at the area assigned in their respective schedules and used ViewRanger® to guide them to each street-point. The surveyors attempted to recruit just one respondent at each street-point, by stopping the first passer-by they saw and inviting them to participate. If the person declined to participate, the surveyors had to wait three minutes before stopping a new passer-by. If a person agreed to participate they were asked to provide informed consent before data collection.

### Data collection and analysis

Informed consent was obtained and data were collected either on-the-spot where privacy could be guaranteed or, if necessary, in a nearby site of the respondent’s choice.

A 35-item structured questionnaire was used; the questionnaire was informed by a previous assessment of communities’ values and preferences for hepatitis C virus self-testing carried out by FIND (the global alliance for diagnostics) [11]. The questionnaire included items on respondents’ socio-demographics; previous experiences with conventional COVID-19 testing; knowledge of other self-test kits; willingness-to-use and -to-pay for a SARS-CoV-2 self-test; barriers to using SARS-CoV-2 self-testing; and likely actions after self-testing for SARS-CoV-2 [12]. This questionnaire was designed in English, translated into Indonesian, and pre-piloted in Jakarta in the premises of the implementing organization, Peduli Hati Bangsa. The finalized questionnaire was developed using the web-based data-collection form builder KoBoToolbox®, re-tested, and deployed in the KoBoCollect® app.

Statistical analyses were conducted using STATA® 14. The primary endpoint of the analysis was willingness to use self-testing in the event they needed a test, as a proxy for the acceptability of self-testing. Associations between respondents’ characteristics, the acceptability of self-testing, and other outcomes of interest were explored, to inform future self-testing distribution programs.

Bivariate and multivariate regression analyses were performed for each outcome of interest, and variables significantly associated with the outcomes at a p-value <0.05 were entered into a multivariate regression model. A logistic regression model was used to identify associations between likelihood to self-test, willingness to pay, and potential predictors. Ordinary least squares (OLS) regression was used for the perception of utility index constructed for appropriate actions taken after testing positive. For the OLS, the outcome on appropriate action taken after testing positive was an index constructed by combining favorable responses for appropriate action taken in relation to face-mask use, self-isolation, communication of a positive result and, warning close contacts.

### Ethics considerations

All respondents provided informed consent to participate. Respondents received no incentive, other than a small bag containing face masks and hand sanitizer. The survey protocol received ethical clearance from the Universitas Katolik Indonesia Atma Jaya (Ref.: 0674A/III/LPPM-PM.10.05/06/2021)

## RESULTS

### Participants’ characteristics

There was a total of 630 respondents, 210 each in Jakarta, Baten, and North Sulawesi (Table 1). Of these, 318 were female. The median age of respondents was 36 (standard deviation (SD) 12.542) years, with just 10.63% of the sample aged ≥56 years. All respondents were born in Indonesia, with Java (*n*=103, 16.35%), Minahasa (*n*=145, 23.02%), and Sunda (*n*=208, 33.02%) being the ethnicities most represented in the sample.

**Table 1.** Respondents’ age, education, and employment status, by sex and location

|  | Rural<br>(Banten, North Sulawesi) |  |  |  | Urban<br>(Jakarta) |  | Sub-total<br>(Rural and urban) |  | Total<br>(N=630) |
| --- | --- | --- | --- | --- | --- | --- | --- | --- | --- |
|  | Banten |  | North Sulawesi |  | Jakarta |  | Rural & Urban |  |  |
|  | Female<br>(n=106) | Male<br>(n=104) | Female<br>(n=109) | Male<br>(n=101) | Female<br>(n=103) | Male<br>(n=107) | Female<br>(n=318) | Male<br>(n=312) |  |
| Mean age in years<br>(standard deviation) | 38.53<br>(12.37) | 35.66<br>(11.90) | 41.14<br>(13.89) | 42.71<br>(14.65) | 38.19<br>(11.73) | 37.69<br>(10.99) | 39.31<br>(12.75) | 38.64<br>(12.87) | 37.41<br>(12.54) |
| Age range |  |  |  |  |  |  |  |  |  |
| 18–35 | 44 (41.51) | 61 (58.65) | 41 (37.61) | 34 (33.66) | 44 (42.72) | 45 (42.06) | 129 (40.57) | 140 (44.87) | 269 (42.70) |
| 36–55 | 53 (50.00) | 35 (33.65) | 55 (50.46) | 45 (44.55) | 53 (51.46) | 53 (49.53) | 161 (50.63) | 133 (42.63) | 294 (46.67) |
| ≥56 | 9 (8.49) | 8 (7.69) | 13 (11.93) | 22 (21.78) | 6 (5.83) | 9 (8.41) | 28 (8.81) | 39 (12.50) | 67 (10.63) |
| Education |  |  |  |  |  |  |  |  |  |
| None | 2 (1.89) | 0(0.00) | 0 (0.00) | 0(0.00) | 3 (2.91) | 0 (0.00) | 5 (1.57) | 0 (0.00) | 5 (0.79) |
| Primary | 38 (35.85) | 23(22.12) | 12 (11.01) | 7(6.93) | 21 (20.39) | 13 (12.15) | 71 (22.33) | 43 (13.78) | 114 (18.10) |
| Secondary | 60 (56.85) | 66(63.46) | 81 (74.31) | 79(78.22) | 56 (54.37) | 66 (61.68) | 197 (61.95) | 211 (67.63) | 408 (64.76) |
| College/Vocational | 4 (3.77) | 8(7.69) | 3 (2.75) | 3(2.97) | 8 (7.77) | 8 (7.48) | 15 (4.72) | 19 (6.09) | 34 (5.40) |
| Degree | 2 (1.89) | 5(4.81) | 12 (11.01) | 11(10.89) | 12 (11.65) | 18 (16.82) | 26 (8.18) | 34 (10.90) | 60 (9.52) |
| Post-graduate | 0 (0.00) | 1(0.96) | 1 (0.92) | 1(0.99) | 0 (0.00) | 2 (1.87) | 1 (0.31) | 4 (1.28) | 5 (0.79) |
| Quranic education | 0 (0.00) | 0(0.00) | 0 (0.00) | 0(0.00) | 1 (0.97) | 0 (0.00) | 1 (0.31) | 1 (0.32) | 2 (0.32) |
| Other | 0 (0.00) | 1(0.96) | 0 (0.00) | 0(0.00) | 2 (1.94) | 0 (0.00) | 2 (0.63) | 0 (0.00) | 2 (0.32) |
| Employment Status |  |  |  |  |  |  |  |  |  |
| Unemployed | 10 (9.43) | 18 (17.31) | 8 (7.34) | 6(7.23) | 5 (4.85) | 5 (4.67) | 23 (7.23) | 29 (9.29) | 52 (8.25) |
| Student | 47 (44.34) | 0 (0.00) | 59 (54.13) | 0(39.62) | 20 (19.42) | 0 (0.00) | 126 (39.62) | 0 (0.00) | 126 (20.00) |
| Employed, part-time | 3 (2.83) | 27 (25.96) | 9 (8.26) | 46(9.75) | 19 (18.45) | 31 (28.97) | 31 (9.75) | 104 (33.33) | 135 (21.43) |
| Employed, full-time | 12 (11.32) | 29 (27.88) | 6 (5.50) | 14(11.95) | 20 (19.42) | 52 (48.60) | 38 (11.95) | 95 (30.45) | 133 (21.11) |
| Self-employed, part-time | 9 (8.49) | 8 (7.69) | 13 (11.93) | 15(11.64) | 15 (14.56) | 9 (8.41) | 37 (11.64) | 32 (10.26) | 69 (10.95) |
| Self-employed, full-time | 25 (23.58) | 22 (21.15) | 14 (12.84) | 15(19.50) | 23 (22.33) | 10 (9.35) | 62 (19.50) | 47 (15.06) | 109 (17.30) |
| Retired, on a pension | 0 (0.00) | 0 (0.00) | 0 (0.00) | 5(0.31) | 1 (0.97) | 0 (0.00) | 1 (0.31) | 5 (1.60) | 6 (0.95) |

Most respondents (*n*=408, 64.76%) had completed secondary school. Completion of university studies varied from as high as 16.82% (*n*=18/107) among males in Jakarta, to 6.51% (*n*=14/215) among females in the provinces. More than two thirds of the sample were employed full-time (*n*=242, 38.41%) or part-time (*n*=204, 32.38%). The largest proportion of unemployed persons was found among male respondents in the rural geographies (*n*=24/205, 11.71%), with the lowest among male respondents in Jakarta (*n*=5/107, 4.67%).

### Experience with COVID-19 testing

Urban respondents reported that they felt more at high- and moderate-risk of COVID-19 than rural respondents (*n*=113/210, 53.80% vs. *n*=98/420, 23.33%) (Table 2). Thirty-four respondents (26 of them from Jakarta) reported that they had COVID-19. Of these, 20 had the disease confirmed by a professional test. There were 68, 79, and 133 respondents, respectively, who perceived that they were living with people with chronic disease (10.79% of the sample), children (12.53%), or elders (21.11%) at increased risk of COVID-19.

**Table 2.**
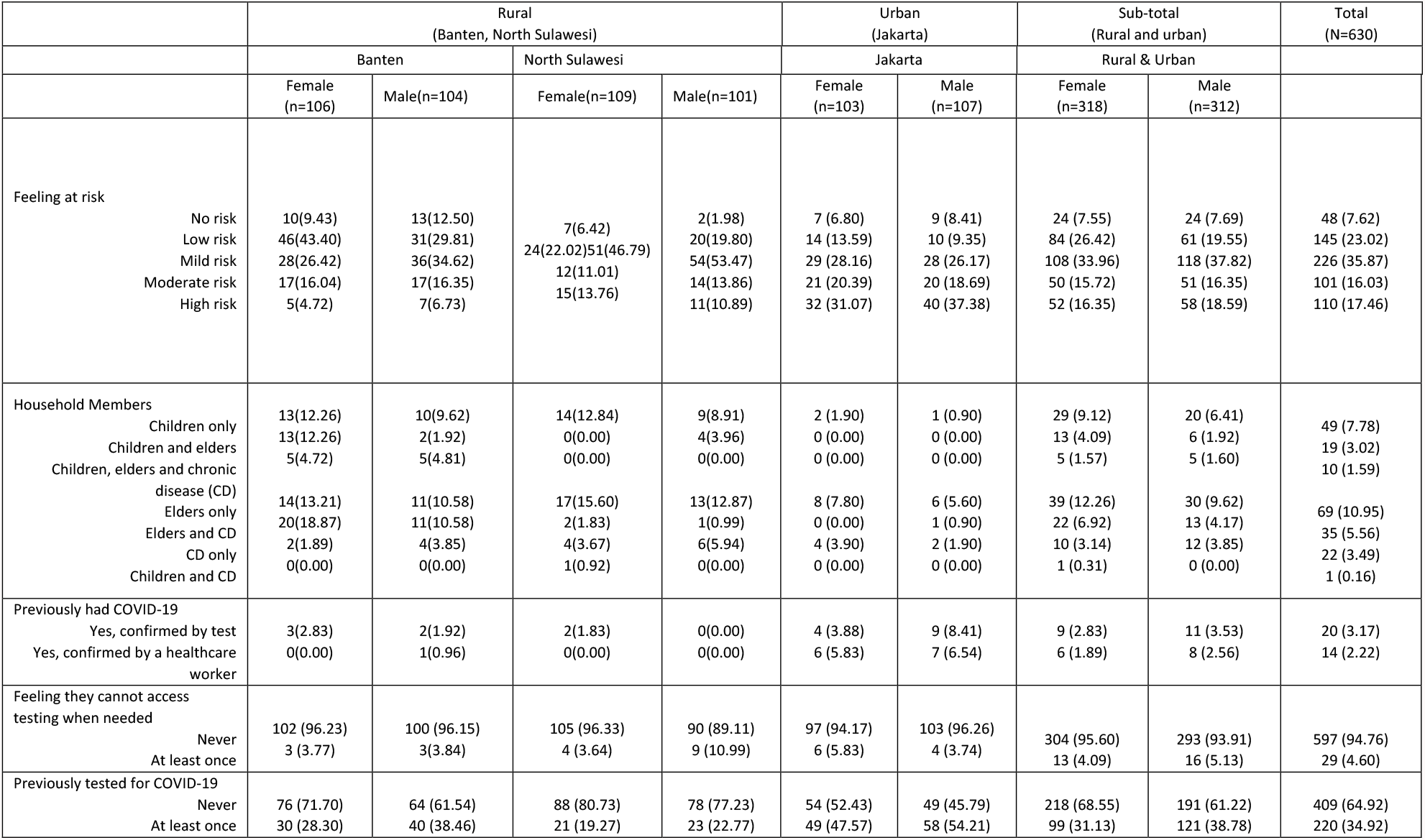
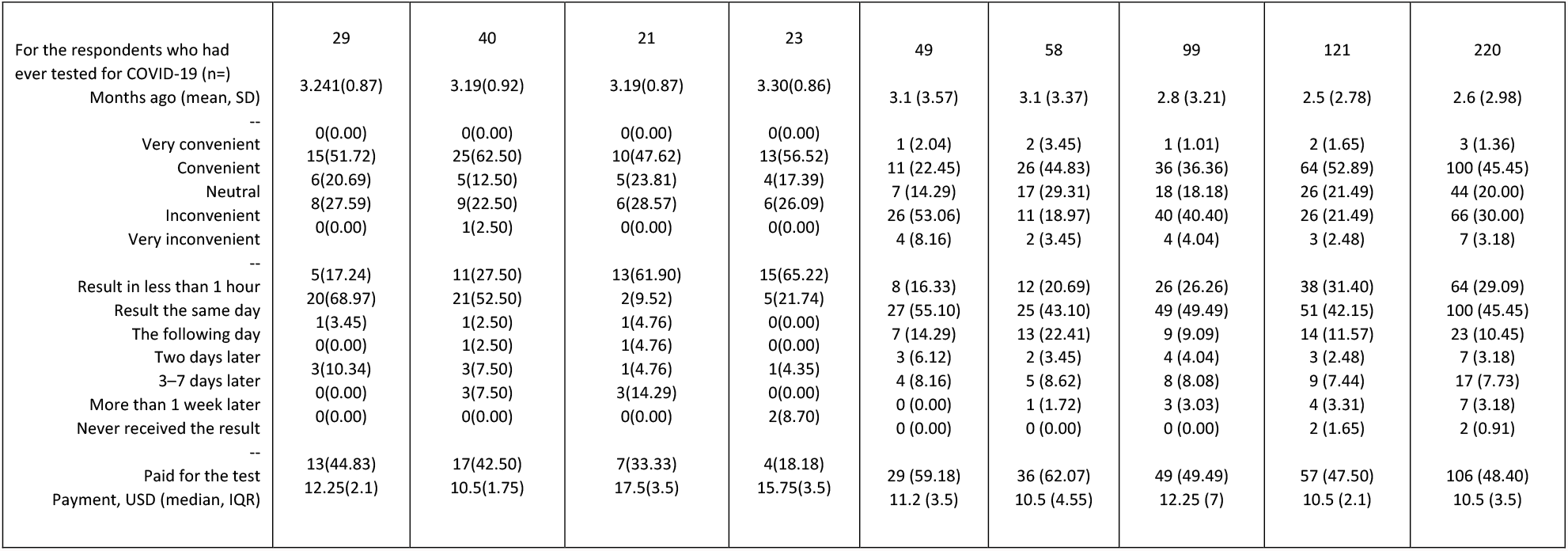
Respondents’ perceived access to and utilization of COVID-19 testing

More than two thirds of rural respondents (*n*=306/420, 74.82%) and almost half of urban respondents (*n*=103/210, 49.05%) had never been tested for COVID-19. Among those who had ever been tested (*n*=219/630, 34.76%), the most recent test was an average of 2.64 (SD 2.98) months ago. Of these, almost half (*n*=103/219, 46.81%) rated their experience as convenient or very convenient; 164 (74.54%) received their test result the same day; 113 (51.60%) did not pay for their test, and the remainder paid a median of 10.5 USD (IQR 3.5USD).

Regarding perceived ease of access to testing, 304 (95.60%) female and 293 (93.91%) male respondents stated they never felt as if they needed a test but could not access it. Of these, 204 (34.17%) reported having ever received a COVID-19 test.

### Acceptability of COVID-19 self-testing

When queried for their awareness of self-testing devices, pregnancy tests were mentioned by 81.65% (*n*=258/318) of females and 61.05% (*n*=185/312) of males. Knowledge of self-testing devices for other infectious diseases was scarce, with devices for HIV (0.86%), malaria (0.15%), and syphilis (0.15%) being the most mentioned. Nevertheless, 15.53% (*n*=96) of the sample mentioned COVID-19 self-testing (Table 3). The highest and lowest rates of knowledge of self-testing for COVID-19 were among females in Jakarta and in the rural areas, respectively (*n*=32/103, 31.10% vs. *n*=9/215, 4.24%).

**Table 3.** Acceptability of self-testing for COVID-19 disease

|  | Rural<br>(Banten, North Sulawesi) |  |  |  | Urban<br>(Jakarta) |  | Sub-total<br>(Rural and urban) |  | Total<br>(N=630) |
| --- | --- | --- | --- | --- | --- | --- | --- | --- | --- |
|  | Banten |  | North Sulawesi |  | Jakarta |  | Rural & Urban |  |  |
|  | Female<br>(n=106)) | Male<br>(n=104) | Female<br>(n=109) | Male<br>(n=101) | Female<br>(n=103) | Male<br>(n=107) | Female<br>(n=318) | Male<br>(n=312) |  |
| Previous knowledge<br>of COVID-19 self-testing | 9 (4.24) | 14 | 0 (0.00) | 0 (0.00) | 32<br>(31.10) | 33 (30.80) | 41 (12.99) | 55 (18.17) | 96 (15.53) |
| Agreement with the<br>concept of COVID-19 self-testing | 49 (46.23) | 61<br>(58.65) | 94<br>(86.24) | 81<br>(80.20) | 48<br>(46.60) | 62 (57.94) | 191 (60.06) | 204 (65.38) | 395 (62.70) |
| Likelihood of using<br>self-testing |  |  |  |  |  |  |  |  |  |
| Very unlikely | 3 (2.83) | 3 (2.88) | 2 (1.83) | 0 (0.00) | 4 (3.88) | 3 (2.80) | 10 (3.14) | 6 (1.92) | 16 (2.54) |
| Unlikely | 40 (37.74) | 31 (29.81) | 6 (5.50) | 3 (2.97) | 28 (27.18) | 21 (19.63) | 74 (23.27) | 55 (17.63) | 129 (20.48) |
| Neutral | 13 12.26) | 8 (7.69) | 12 (11.01) | 18 (17.82) | 27 (26.21) | 25 (23.36) | 52 (16.35) | 51 (16.35) | 103 (16.35) |
| Likely | 45 (42.45) | 56 (53.85) | 73 (66.97) | 63 (62.38) | 30 (29.13) | 39 (36.45) | 148 (46.54) | 158 (50.64) | 306 (48.57) |
| Very likely | 5 (4.72) | 6 (5.77) | 16 (14.68) | 17 (16.83) | 14 (13.59) | 19 (17.76) | 35 (11.01) | 42 (13.46) | 77 (12.22) |
| --- | --- | --- | --- | --- | --- | --- | --- | --- | --- |
| Likelihood (mean,<br>SD) | 3.084<br>(1.05) | 3.29<br>(1.05) | 3.87<br>(0.79) | 3.93<br>(0.68) | 3.21<br>(1.108) | 3.47<br>(1.0843) | 3.39<br>(1.047) | 3.56<br>(.9932) | 3.48<br>(1.023) |
| Willing to serially<br>test |  |  |  |  |  |  |  |  |  |
| Yes | 43 (40.57) | 47 (45.19) | 77 (70.64) | 79(78.22) | 65 (63.10) | 72 (67.30) | 185 (58.20) | 198 (63.50) | 383 (60.82) |
| No | 44 (41.51) | 44 (42.13) | 24 (22.02) | 11(10.89) | 33 (32.04) | 28 (26.17) | 101 (31.76) | 83 (26.60) | 184 (29.21) |
| For the respondents<br>willing to pay for a<br>self-test (n=) | 79 (74.53) | 75 (72.12) | 31 (28.44) | 29 (28.71) | 86 (83.49) | 91 (85.04) | 196 (61.63) | 195 (62.50) | 391 (62.06) |
| Maximum<br>acceptable payment<br>in USD (median,<br>IQR) | 1.05<br>(1.05) | 1.4<br>(0.7) | 1.75<br>(2.45) | 2.45<br>(2.1) | 3.5<br>(3.5) | 2.1<br>(2.1) | 1.4<br>(2.55) | 1.75<br>(2.45) | 1.4<br>(2.45) |

Almost two in three respondents (*n*=395, 62.70%) agreed with the concept of people being able to self-test at home for COVID-19. Agreement with the concept of self-testing was slightly lower in Jakarta than in the rural geographies for both females (57.94% vs. 69.27%, respectively) and males (46.60% vs. 66.51%, respectively).

If freely available and recommended by health authorities, 383 (60.82%) respondents were willing to test at least once per week. The likelihood to use a self-test for COVID-19 when needed rated highly, with an average rating of 3.48/5 (SD 1.023) in total, and 3.39/5 (SD 1.047) for females and 3.56/5 (SD 0.9932) for males. Overall, if COVID-19 self-testing were available in Indonesia, 77 (12.22%) and 306 (48.57%) respondents would be very likely or likely, respectively, to use them if they felt it necessary; of these, the majority (*n*=281/383, 73.36%) were from the rural geographies. While only 42.72% (*n*=44/103) of females from Jakarta answered “very likely” or “likely” to use self-testing, 69.27% (*n*=142/205) of males in rural areas gave these responses.

As per the bivariate analyses (Table 5), potential predictive factors of likelihood to use self-testing included living in a rural area, knowledge of pregnancy self-tests, feeling at mild risk of COVID-19, agreement with the concept of self-testing for COVID-19, being employed part-time, and having completed any education above primary school. The multivariate model showed that rural respondents (adjusted odds ratio (AOR): 3.26, confidence interval (95%CI): 2.16–4.93, p<0.001), having secondary education (AOR: 1.78, CI: 1.15–2.79, p<0.01) or a college degree (AOR: 3.9, CI: 2.08– 7.31, p<0.001), and those who were working part-time (AOR: 1.95, CI: 1.21–3.15, p<0.006) had a comparatively higher odds of using self-testing kits.

If not available free-of-charge, 391 (62.06%) respondents stated that they would be willing to pay for a self-testing device if they needed it (i.e., a median of 1.4 USD (IQR 2.45) (Table 3). The bivariate associations showed that respondent characteristics such as urban location, age <36 years, having secondary education or a college degree, in full-time employment, and having higher perception of risk of COVID-19 were potential predictors of willingness to pay for self-testing devices (Table 5). The multivariate model confirmed that individuals aged 36–55 years (AOR: 0.56, CI: 0.38–0.82, p<0.003) and >56 years (AOR: 0.025, CI:0.13–0.047, p<0.001) were less likely to pay for a self-testing device compared with individuals aged <36 years. Rural residents were less likely to pay for self-testing kits than urban residents (AOR: 0.23. CI:0.147–0.36, p<0.001). The respondents with a moderate to high perception of COVID-19 risk were more likely to pay for a self-test (AOR: 1.99, CI:1.31–3.03, p<0.001).

### Actions upon self-testing for COVID-19

Most respondents stated that if they performed a SARS-CoV-2 self-test and its result was positive, they would communicate the result to the relevant authorities (*n*=542, 86.03%), visit a health facility to request post-test counseling (*n*=509, 80.79%), self-isolate (*n*=614, 97.46%), and warn their contacts (*n*=570, 90.48%) (Table 4).

**Table 4.** Actions following a SARS-CoV-2 self-test

|  | Rural<br>(Banten, North Sulawesi) |  |  |  | Urban<br>(Jakarta) |  | Sub-total<br>(Rural and urban) |  | Total<br>(N=630) |
| --- | --- | --- | --- | --- | --- | --- | --- | --- | --- |
|  | Banten |  | North Sulawesi |  | Jakarta |  | Rural & Urban |  |  |
|  | Female<br>(n=106) | Male<br>(n=104) | Female<br>(n=109) | Male<br>(n=101) | Female<br>(n=103) | Male<br>(n=107) | Female<br>(n=318) | Male<br>(n=312) |  |
| Practices following receipt of a positive self-test result |  |  |  |  |  |  |  |  |  |
| Communicate the result to a clinic, hospital, and/or COVID hotline | 87<br>(82.08) | 88(84.62) | 86<br>(78.90) | 84(83.17) | 97<br>(94.17) | 100<br>(93.46) | 270<br>(84.91) | 272<br>(87.18) | 542<br>(86.03) |
| Go in person to a clinic or hospital to get post-test counseling from a healthcare worker | 76<br>(71.70) | 73(70.19) | 86<br>(78.90) | 91(90.10) | 90<br>(87.38) | 93<br>(86.92) | 252<br>(79.25) | 257<br>(82.37) | 509<br>(80.79) |
| Self-isolate | 105<br>(99.06) | 99(95.19) | 103<br>(94.50) | 99(98.02) | 102<br>(99.03) | 106<br>(99.07) | 310<br>(97.48) | 304<br>(97.44) | 614<br>(97.46) |
| Identify and warn close contacts | 88<br>(83.02) | 92(88.46) | 90<br>(82.57) | 95(94.06) | 102<br>(99.03) | 103<br>(96.26) | 280<br>(88.05) | 290<br>(92.95) | 570<br>(90.48) |
| Inform their employer (n= respondents employed) | n=49<br>28<br>(26.42)) | n=86<br>63(60.58) | n=42<br>34<br>(31.19) | n=90<br>80(79.21) | n=77<br>52<br>(67.53) | n=102<br>79<br>(77.45) | n=168<br>114<br>(67.86) | n=278<br>222<br>(79.86) | n=446<br>336<br>75.34 |
| Practices following receipt of a negative self-test for a person with symptoms and exposed to a COVID-19 case |  |  |  |  |  |  |  |  |  |
| Stop self-isolation | 82<br>(77.36) | 77(74.04) | 76<br>(69.72) | 75(74.26) | 76<br>(73.79) | 84<br>(78.50) | 234<br>(73.58) | 236<br>(75.64) | 470<br>(74.60) |
| Stop wearing a face-mask | 11<br>(10.38) | 8(7.69) | 3(2.75) | 2(1.98) | 2 (1.94) | 6 (5.61) | 16 (5.03) | 16 (5.13) | 32 (5.08) |
| Stop social distancing | 31<br>(29.25) | 25(24.04) | 10 (9.17) | 8 (7.92) | 4 (3.88) | 9 (8.41) | 45<br>(14.15) | 42 (13.46) | 87<br>(13.81) |

**Table 5:** Bivariate and multivariate logistic regression analysis depicting the association between independent variables and likeliness to test and willingness to pay for COVID-19 self-testing kits

|  | Bivariate |  |  |  |  |  |  | Multivariate |  |
| --- | --- | --- | --- | --- | --- | --- | --- | --- | --- |
|  | (1) | (2) | (3) | (4) | (5) | (6) | (7) | (8) | (9) |
| Variable | Likely to use a self-test<br>(odds ratio (standard error)) |  |  |  |  |  |  | Likely to use a self-test (with controls)<br>(adjusted odds ratio (standard error)) | p-value |
| Rural | 2.140***<br>(0.370) |  |  |  |  |  |  | 3.263***<br>(0.686) | <0.001 |
| Female |  | 0.759*<br>(0.124) |  |  |  |  |  | 1.015<br>(0.196) | 0.940 |
| Aged 36–55 years |  |  | 1.027<br>(0.178) |  |  |  |  |  |  |
| Aged ≥56 years |  |  | 0.963<br>(0.269) |  |  |  |  |  |  |
| Secondary education |  |  |  | 1.898***<br>(0.406) |  |  |  | 1.788**<br>(0.405) | 0.010 |
| College degree or higher |  |  |  | 3.231***<br>(0.956) |  |  |  | 3.903***<br>(1.250) | <0.001 |
| Other education (Quranic or none) |  |  |  | 0.575<br>(0.421) |  |  |  | 0.625<br>(0.499) | 0.556 |
| Part-time employment |  |  |  |  | 1.803***<br>(0.385) |  |  | 1.949***<br>(0.478) | 0.006 |
| Full-time employment |  |  |  |  | 0.987<br>(0.195) |  |  | 1.085<br>(0.252) | 0.726 |
| High Perception of risk of COVID-19 |  |  |  |  |  | 0.907<br>(0.156) |  |  |  |
| Higher household risk |  |  |  |  |  |  | 0.684**<br>(0.118) | 0.473***<br>(0.0957) | <0.001 |
| Mean | 0.944 | 1.786 | 1.538 | 0.869 | 1.300 | 1.602 | 1.760 |  |  |
| Observations | 630 | 630 | 630 | 630 | 630 | 630 | 630 | 630 |  |
| Wald chi-square | 19.39 | 2.830 | 0.0608 | 19.16 | 10.81 | 0.320 | 4.809 | 57.02 |  |
| p-value | <0.001 | 0.0925 | 0.970 | <0.001 | 0.00449 | 0.572 | 0.0283 | <0.001 |  |
| Adj R<sup>2</sup> | 0.0232 | 0.00337 | 0 | 0.0237 | 0.0133 | 0.000379 | 0.00569 | 0.0829 |  |
| Variable | Willingness to pay (odds ratio (standard error)) |  |  |  |  |  |  | Willingness to pay (with controls)<br>(adjusted odds ratio (standard error)) | p-value |
| Rural | 0.194***<br>(0.0413) |  |  |  |  |  |  | 0.231***<br>(0.0533) | <0.001 |
| Female |  | 0.964<br>(0.158) |  |  |  |  |  |  |  |
| Aged 36–55 years |  |  | 0.638**<br>(0.115) |  |  |  |  | 0.558***<br>(0.111) | 0.003 |
| Aged ≥56 years |  |  | 0.217***<br>(0.0631) |  |  |  |  | 0.248***<br>(0.0805) | <0.001 |
| Secondary education |  |  |  | 1.649**<br>(0.352) |  |  |  | 1.329<br>(0.341) | 0.268 |
| College degree or higher |  |  |  | 2.960***<br>(0.882) |  |  |  | 1.761*<br>(0.568) | 0.080 |
| Other education (Quranic or none) |  |  |  | 2.000<br>(1.464) |  |  |  | 1.308<br>(1.161) | 0.762 |
| Part-time employment |  |  |  |  | 1.150<br>(0.236) |  |  | 0.740<br>(0.171) | 0.192 |
| Full-time employment |  |  |  |  | 1.979***<br>(0.405) |  |  | 1.175<br>(0.276) | 0.492 |
| High Perception of risk of COVID-19 |  |  |  |  |  | 3.063***<br>(0.593) |  | 1.992***<br>(0.429) | 0.001 |
| Higher household risk |  |  |  |  |  |  | 0.802<br>(0.140) |  |  |
| Mean | 5.364 | 1.667 | 2.405 | 1 | 1.217 | 1.171 | 1.760 |  |  |
| Observations | 630 | 630 | 630 | 630 | 630 | 630 | 630 | 630 |  |
| Wald chi-square | 59.16 | 0.0499 | 28.19 | 13.52 | 12.66 | 33.45 | 1.601 | 87.94 |  |
| p-value | 0 | 0.823 | <0.001 | 0.00364 | 0.00179 | <0.001 | 0.206 | <0.001 |  |
| Adj R2 | 0.0856 | 0 | 0.0359 | 0.0169 | 0.0156 | 0.0440 | 0.00191 | 0.144 |  |

Table notes: Robust standard errors in parentheses \*\*\* p<0.01, \*\* p<0.05, \* p<0.1. The omitted/comparison category for each of the covariates were: Urban for rural, male for female, 18–34 years for age, unemployed/student/retired on pension for employment, primary education for education, lower perception of risk/no risk for high perception of risk and households that did not have any perceived at-risk individuals i.e., children, old people, and people with chronic diseases. Likely to use a self-test was a binary variable constructed using those who responded “Likely/Very likely” in the survey on their likelihood of using a COVID-19 self-test if available. Willingness to pay was a binary variable constructed using those who responded any value other than “0” in the survey on how much were they willing to pay for a COVID-19 self-test. People with a higher perception of risk of COVID-19 were those who reported moderate- to high-risk perception. Higher-risk households were those with children, elderly, and members with chronic diseases. The multivariate analyses included controls for gender, age, ethnicity, location (rural/urban), employment, perception of risk, and higher household risk.

Respondents’ preferred channels for reporting positive results were attending a clinic in person (*n*=509, 81.05%) and use of community healthcare workers (*n*=383, 60.99%). Just 41 (6.53%) respondents stated they would not report a positive result.

In the event that an individual had symptoms and knew they had been exposed to a person with COVID-19 but their result was negative, the majority would not stop social distancing (only *n*=87, 13.81% would) or stop wearing masks (only *n*=32, 5.08% would). However, three in four respondents would stop self-isolating (*n*=470, 74.60%). A significant association with agreement with the concept of self-testing and being very likely or likely to using a self-testing if available in Indonesia was found with communicating a positive result, seeking post-test counseling, informing an employer, identifying close contacts, and with not stopping self-isolation, wearing masks, or social distancing following a negative result.

The bivariate associations showed that respondents’ characteristics such as urban location, being male, having a college degree or higher education, part-time or full-time employment, higher perception of COVID-19 risk, and high-risk household members were strong predictors of the index of actions taken following a positive COVID-19 self-test (Table 6). The multivariate analysis confirmed that respondents from rural areas were 0.28 SD less likely to fare well on the afore-mentioned index than those from urban areas (95%CI: -0.43 to -0.12, p<0.001). Similarly, part-time employed were 0.19 SD (90%CI: -0.18 to 0.41, p<0.10) more likely to take appropriate action after a positive self-test in comparison to those where were unemployed (includes individuals that were unemployed, students and retired).

**Table 6:**
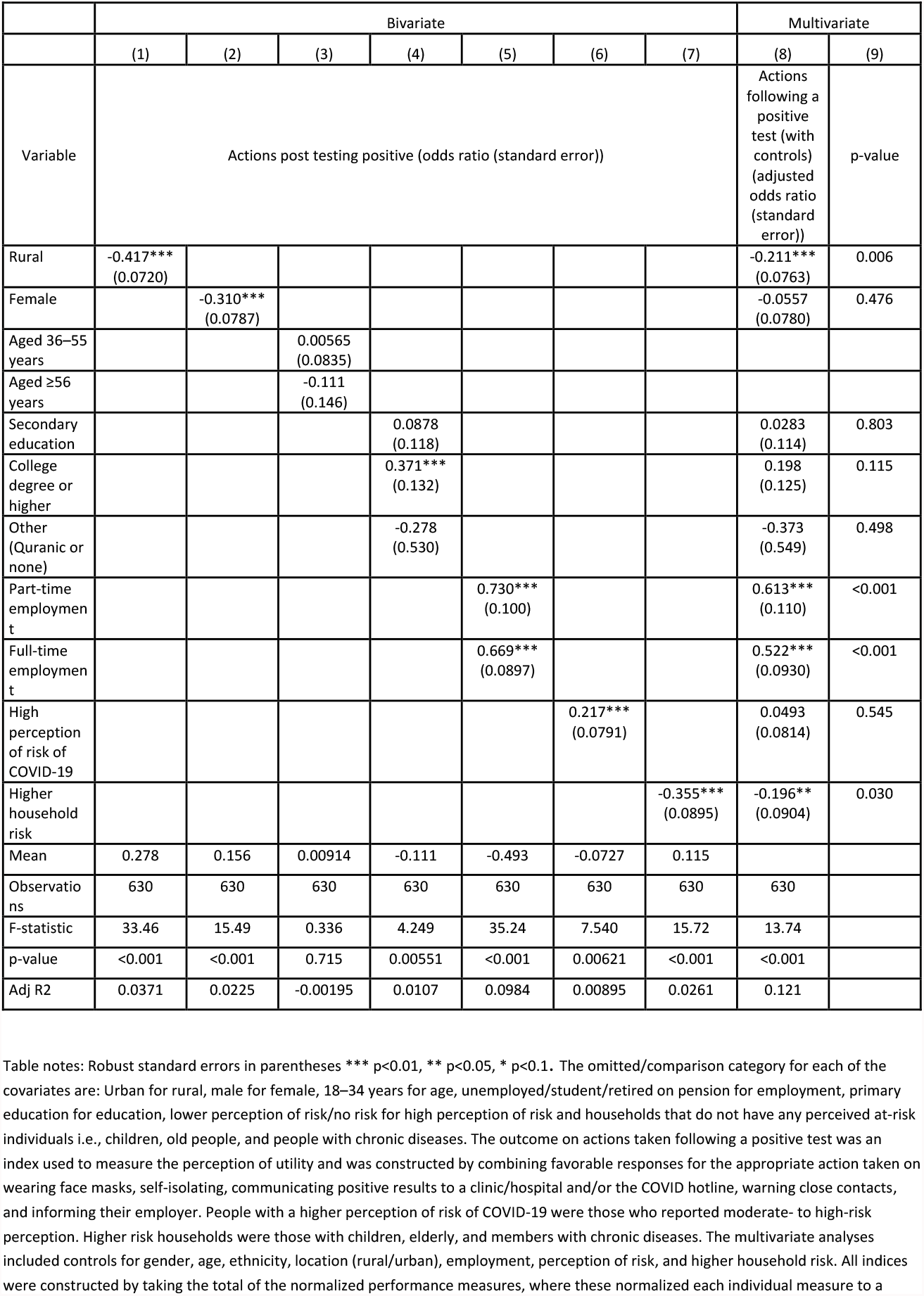
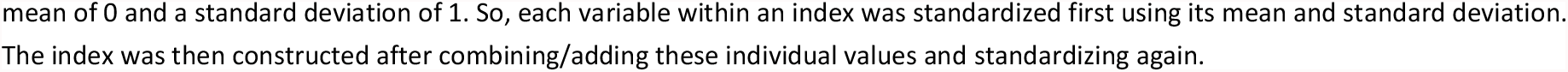
Bivariate and multivariate ordinary least squares (OLS) regression analysis depicting the association between independent variables and actions taken following a positive result using a COVID-19 self-testing kit

|  | Bivariate |  |  |  |  |  |  | Multivariate |  |
| --- | --- | --- | --- | --- | --- | --- | --- | --- | --- |
|  | (1) | (2) | (3) | (4) | (5) | (6) | (7) | (8) | (9) |
| Variable | Actions post testing positive (odds ratio (standard error)) |  |  |  |  |  |  | Actions following a positive test (with controls) (adjusted odds ratio (standard error)) | p-value |
| Rural | -0.417***<br>(0.0720) |  |  |  |  |  |  | -0.211***<br>(0.0763) | 0.006 |
| Female |  | -0.310***<br>(0.0787) |  |  |  |  |  | -0.0557<br>(0.0780) | 0.476 |
| Aged 36–55 years |  |  | 0.00565<br>(0.0835) |  |  |  |  |  |  |
| Aged ≥56 years |  |  | -0.111<br>(0.146) |  |  |  |  |  |  |
| Secondary education |  |  |  | 0.0878<br>(0.118) |  |  |  | 0.0283<br>(0.114) | 0.803 |
| College degree or higher |  |  |  | 0.371***<br>(0.132) |  |  |  | 0.198<br>(0.125) | 0.115 |
| Other (Quranic or none) |  |  |  | -0.278<br>(0.530) |  |  |  | -0.373<br>(0.549) | 0.498 |
| Part-time employment |  |  |  |  | 0.730***<br>(0.100) |  |  | 0.613***<br>(0.110) | <0.001 |
| Full-time employment |  |  |  |  | 0.669***<br>(0.0897) |  |  | 0.522***<br>(0.0930) | <0.001 |
| High perception of risk of COVID-19 |  |  |  |  |  | 0.217***<br>(0.0791) |  | 0.0493<br>(0.0814) | 0.545 |
| Higher household risk |  |  |  |  |  |  | -0.355***<br>(0.0895) | -0.196**<br>(0.0904) | 0.030 |
| Mean | 0.278 | 0.156 | 0.00914 | -0.111 | -0.493 | -0.0727 | 0.115 |  |  |
| Observations | 630 | 630 | 630 | 630 | 630 | 630 | 630 | 630 |  |
| F-statistic | 33.46 | 15.49 | 0.336 | 4.249 | 35.24 | 7.540 | 15.72 | 13.74 |  |
| p-value | <0.001 | <0.001 | 0.715 | 0.00551 | <0.001 | 0.00621 | <0.001 | <0.001 |  |
| Adj R2 | 0.0371 | 0.0225 | -0.00195 | 0.0107 | 0.0984 | 0.00895 | 0.0261 | 0.121 |  |

Table notes: Robust standard errors in parentheses \*\*\* p<0.01, \*\* p<0.05, \* p<0.1. The omitted/comparison category for each of the covariates are: Urban for rural, male for female, 18–34 years for age, unemployed/student/retired on pension for employment, primary education for education, lower perception of risk/no risk for high perception of risk and households that do not have any perceived at-risk individuals i.e., children, old people, and people with chronic diseases. The outcome on actions taken following a positive test was an index used to measure the perception of utility and was constructed by combining favorable responses for the appropriate action taken on wearing face masks, self-isolating, communicating positive results to a clinic/hospital and/or the COVID hotline, warning close contacts, and informing their employer. People with a higher perception of risk of COVID-19 were those who reported moderate- to high-risk perception. Higher risk households were those with children, elderly, and members with chronic diseases. The multivariate analyses included controls for gender, age, ethnicity, location (rural/urban), employment, perception of risk, and higher household risk. All indices were constructed by taking the total of the normalized performance measures, where these normalized each individual measure to a
mean of 0 and a standard deviation of 1. So, each variable within an index was standardized first using its mean and standard deviation. The index was then constructed after combining/adding these individual values and standardizing again.

## DISCUSSION

SARS-CoV-2 self-testing represents an innovative technology that could positively impact case detection in Indonesia. This survey, which involved 630 individuals from a variety of urban and rural geographies in the country, suggested that the Indonesian public would be willing to use a self-test for COVID-19 and would react positively if they received a positive result. Two in three respondents expressed agreement with the concept of home self-testing for COVID-19 and stated that they would use them if they felt they needed to test. The majority of respondents expressed that, in accordance with health authorities’ main recommendations [13, 14], they would report a positive result, request post-test counseling, self-isolate, and notify their close contacts.

These results must be interpreted with caution. The fact that one third of respondents did not express interest in using a SARS-CoV-2 self-test might not be related to them disagreeing with the innovation *per se*, but rather to them having convenient access to conventional COVID-19 testing in Indonesia or to their self-perception of being at low risk of experiencing severe COVID-19 disease. It is also worth reflecting on the fact that, in both urban and rural geographies, females were less likely than males to state that they would use self-testing. In Indonesia, women exhibit an increased likelihood compared with men to attend health facilities [15, 16], and this health-seeking behavior may extend to women feeling more comfortable than men in visiting health facilities to request conventional COVID-19 testing. Hence, females may not place as much value as males on the option of confidential testing in private, without the assistance of a healthcare worker. Further qualitative research may help to clarify the reasons for these differences once self-testing devices become widely available in Indonesia.

There are concerns that people self-testing for infectious diseases may not behave in the optimal manner to maximize public health benefits of the self-test upon receiving their result. These concerns have been assessed in HIV and hepatitis C self-testing acceptability studies [17, 18]. In the present survey, the majority of respondents stated that, if they self-tested for SARS-CoV-2, they would continue adhering to health authority recommendations such as wearing masks and/or reporting the result. This finding is aligned with the results of other SARS-CoV-2 self-testing studies carried out in the United Kingdom [19] and Germany [10]. However, in our survey, three in four respondents said they would not isolate following a negative self-test even if they had COVID-19 symptoms and they knew that they had been in contact with a COVID-19 patient. While the need to ensure they can generate income to provide for their offspring and themselves is a reason to not isolate, this survey did not assess the reasons behind participants’ responses. It might be possible that they would not isolate but they would still report a negative result and request a confirmatory test at their nearest facility. As there is potential for social harm arising from the risk that self-test users with COVID-19-compatible symptoms do not isolate, it is recommended that the distribution of self-tests in Indonesia be performed in conjunction with clear sensitization on what actions to take following a positive or a negative result.

Respondents in our survey would prefer to report self-test results directly to a healthcare worker in a clinic or hospital or through community health workers. Despite the public’s distrust of the government for its management of the COVID-19 pandemic [20], this finding suggests the public do trust healthcare workers. While it is important to develop phone- and web-based reporting mechanisms for self-test users living in remote areas and in the islands, it is also important to capitalize on the good relationships that many Indonesians have established with their healthcare workers.

The favorable attitudes toward self-testing found in our survey might be related to the high level of awareness of self-tests for the diagnosis of non-infectious and infectious diseases, as well as to awareness of the local availability of saliva-based SARS-CoV-2 tests [21, 22]. Major news channels and websites as well as information from telemedicine platforms may have increased awareness of the accessibility of these kits [7, 8]. However, despite the public’s awareness of diagnostics for home use, Indonesia is a highly populated, middle-income country where many households lack the resources to afford new technologies for health. In this regard, it is significant that 62.06% of the sample were willing to pay for a self-test and that the median they would be willing to pay was as 1.4 USD. This finding has major implications for the delivery of SARS-CoV-2 self-testing. For self-testing to have an impact in terms of case detection in Indonesia, quality, affordable SARS-CoV-2 self-tests will need to be brough to market.

The importance of self-testing in low- and middle-income countries where diagnostic testing capacity is scarce has been highlighted previously [23]. Ours is, to the best of our knowledge, the only survey of the Indonesian public’s values in relation to SARS-CoV-2 self-testing, and a direct comparison of our findings with those of similar studies is currently not possible. However, previous studies of HIV self-testing in Indonesia suggest that many people may appreciate SARS-CoV-2 self-testing, for myriad reasons: fear of shame, embarrassment, and social exclusion; issues around breach of confidentiality; fear of invasive testing methods; and concerns around privacy and inconvenience [24, 25].

A number of limitations must be considered. During the implementation of this survey in rural geographies, the surveyors found a significant number of neighborhoods whose perimeters could not be crossed by order of local authorities. The surveyors had to select new, nearby recruitment street-points, which may have introduced recruitment bias. In Jakarta, the surveyors worked on some very crowded streets. Many individuals who were approached (*n*=235) refused to even let the surveyors explain the purpose of the survey; it is impossible to know whether these individuals’ characteristics differed from consenting respondents’ characteristics. Another limitation relates to the country’s cultural and socio-economic diversity. Despite the choice of Banten and North Sulawesi as geographies with very different social strata, it is possible that the survey findings would have been different if other regions had been sampled. Finally, it must be noted that the cross-sectional design limited our capacity to statistically establish causal relationships between likelihood to use self-testing, willingness to pay, and associated factors.

In conclusion, the Indonesian public appreciates self-testing diagnostics for detection of infectious diseases and would use SARS-CoV-2 self-tests if widely available. As recommended by health authorities, it is highly probable that self-test users would report positive results and would self-isolate, warn their contacts, and continue wearing face masks. In Indonesia, self-testing kits should be introduced in a way that would encourage users to access confirmatory testing and COVID-19 treatment following a positive self-test result, as well as continue adhering to preventive behaviors, irrespective of the test result.

## Data Availability

All data produced in the present study are available upon reasonable request to the authors

## References

1. Zhu H, Wei L, Niu P. The novel coronavirus outbreak in Wuhan, China. Glob Health Res Policy. 2020;5:6.

2. Cucinotta D, Vanelli M. WHO Declares COVID-19 a Pandemic. Acta Biomed. 2020;91(1):157–60.

3. Setiati S, Azwar MK. COVID-19 and Indonesia. Acta Med Indones. 2020;52(1):84–9.

4. Johns Hopkins Coronavirus Resource Center. Indonesia: COVID-19 Overview 2021 [Available from: https://coronavirus.jhu.edu/region/indonesia.

5. Hendarwan H, Syachroni S, Aryastami NK, Su’udi A, Susilawati MD, Despitasari M, et al. Assessing the COVID-19 diagnostic laboratory capacity in Indonesia in the early phase of the pandemic. WHO South East Asia J Public Health. 2020;9(2):134–40.

6. Aisyah DN, Mayadewi CA, Igusti G, Manikam L, Adisasmito W, Kozlakidis Z. Laboratory Readiness and Response for SARS-Cov-2 in Indonesia. Front Public Health. 2021;9:705031.

7. Aditya R. QuickSpit: Tes Covid-19 Pakai Air Liur Dinilai Akurat Deteksi Virus Corona: Suara.com; 2021 [Available from: https://www.suara.com/health/2021/07/26/204615/quickspit-tes-COVID-19-pakai-air-liur-dinilai-akurat-deteksi-virus-corona?page=all.

8. Anastasia T. Fakta-fakta tentang Tes COVID-19 Menggunakan Air Liur: klikdokter; 2021 [Available from: https://www.klikdokter.com/info-sehat/read/3648725/fakta-fakta-tentang-tes-covid-19-menggunakan-air-liur.

9. Ganguli I, Bassett IV, Dong KL, Walensky RP. Home testing for HIV infection in resource-limited settings. Curr HIV/AIDS Rep. 2009;6(4):217–23.

10. Wachinger J, Schirmer M, Täuber N, McMahon SA, Denkinger CM. Experiences with opt-in, at-home screening for SARS-CoV-2 at a primary school in Germany: an implementation study. BMJ Paediatr Open. 2021;5(1):e001262.

11. WHO. Recommendations and guidance on hepatitis C virus self-testing: web annex D: values and preferences on hepatitis C virus self-testing. Geneva: World Health Organization; 2021 2021.

12. Shilton S, Ivanova Reipold E, Roca Álvarez A, Martínez-Pérez GZ. Assessing Values and Preferences Toward SARS-CoV-2 Self-testing Among the General Population and Their Representatives, Health Care Personnel, and Decision-Makers: Protocol for a Multicountry Mixed Methods Study. JMIR Res Protoc. 2021;10(11):e33088.

13. WHO. Advice for the public: Coronavirus disease (COVID-19) Geneva: WHO; 2021 [Available from: https://www.who.int/emergencies/diseases/novel-coronavirus-2019/advice-for-public.

14. Better Work Indonesia. Compilation of Guidelines on Covid-19 Transmission, Prevention and Management and the Best Practices in the Workplace 2020 [Available from: https://betterwork.org/wp-content/uploads/2020/04/BWI_covid_guidance_eng_web.pdf

15. Handayani PW, Dartanto T, Moeis FR, Pinem AA, Azzahro F, Hidayanto AN, et al. The regional and referral compliance of online healthcare systems by Indonesia National Health Insurance agency and health-seeking behavior in Indonesia. Heliyon. 2021;7(9):e08068.

16. Ahmad RA, Richardus JH, de Vlas SJ. Care-seeking behaviour among individuals with TB symptoms in Jogjakarta Province, Indonesia: a community-based study. Int Health. 2013;5(1):51–7.

17. Jamil MS, Eshun-Wilson I, Witzel TC, Siegfried N, Figueroa C, Chitembo L, et al. Examining the effects of HIV self-testing compared to standard HIV testing services in the general population: A systematic review and meta-analysis. EClinicalMedicine. 2021;38:100991.

18. Martínez-Pérez GZ, Nikitin DS, Bessonova A, Fajardo E, Bessonov S, Shilton S. Values and preferences for hepatitis C self-testing among people who inject drugs in Kyrgyzstan. BMC Infect Dis. 2021;21(1):609.

19. Wanat M, Logan M, Hirst JA, Vicary C, Lee JJ, Perera R, et al. Perceptions on undertaking regular asymptomatic self-testing for COVID-19 using lateral flow tests: a qualitative study of university students and staff. BMJ Open. 2021;11(9):e053850.

20. Riefky, Hutasoit I, Nopiyanto A, Nugrahani H, Zulkarnain R. Growing public distrust towards the Indonesian Government for lack of response to COVID-19 outbreak. IOP Conference Series: Earth and Environmental Science. 2021;716:012072.

21. Kalbe. Kalbe Launches the First Ever Saliva-Based COVID-19 Test Kit which is the Brainchild of Indonesia’s Own Researchers 2021 [Available from: https://www.kalbe.co.id/news-and-events/ArtMID/443/ArticleID/918/Kalbe-Launches-the-First-Ever-Saliva-Based-COVID-19-Test-Kit-which-is-the-Brainchild-of-Indonesia%E2%80%99s-Own-Researchers.

22. Bio Farma. Bio Farma X Nusantics Launches Limited Bio Saliva, Covid-19 PCR Detection Test With Gargle Method 2021 [Available from: https://www.biofarma.co.id/en/latest-news/detail/bio-farma-x-nusantics-launches-limited-bio-saliva-covid19-pcr-detection-testwith-gargle-method.

23. Boum Y, Eyangoh S, Okomo MC. Beyond COVID-19-will self-sampling and testing become the norm? Lancet Infect Dis. 2021;21(9):1194–5.

24. Wulandari LPL, Ruddick A, Guy R, Kaldor J. “Self-testing sounds more private, rather than going to the clinic and everybody will find out”: Facilitators and barriers regarding HIV testing among men who purchase sex in Bali, Indonesia. PLoS One. 2019;14(4):e0214987.

25. Wulandari LPL, Kaldor J, Guy R. Uptake and acceptability of assisted and unassisted HIV self-testing among men who purchase sex in brothels in Indonesia: a pilot intervention study. BMC Public Health. 2020;20(1):730.

